# A multicenter prospective randomized controlled trial of high sensitivity cardiac troponin I-guided combination angiotensin receptor blockade and beta-blocker therapy to prevent anthracycline cardiotoxicity: the Cardiac CARE trial

**DOI:** 10.1101/2023.02.20.23286206

**Authors:** Peter A. Henriksen, Peter Hall, Iain R. MacPherson, Shruti S Joshi, Trisha Singh, Morag Maclean, Steff Lewis, Aryelly Rodriguez, Alex Fletcher, Russell J Everett, Harriet Stavert, Angus Broom, Lois Eddie, Lorraine Primrose, Heather McVicars, Pam McKay, Annabel Borley, Clare Rowntree, Simon Lord, Graham Collins, John Radford, Amy Guppy, Michelle C Williams, Alan Japp, John R. Payne, David E. Newby, Nick L. Mills, Olga Oikonomidou, Ninian N. Lang

## Abstract

**Background:** Anthracycline-induced cardiotoxicity has a variable incidence, and the development of left ventricular dysfunction is preceded by elevations in cardiac troponin concentrations. Beta-adrenergic receptor blocker and renin-angiotensin-system inhibitor therapies have been associated with modest cardioprotective effects in unselected patients receiving anthracycline chemotherapy.

**Methods:** In a multicenter prospective randomized open label blinded endpoint trial, patients with breast cancer and non-Hodgkin lymphoma receiving anthracycline chemotherapy underwent serial high-sensitivity cardiac troponin testing and cardiac magnetic resonance imaging before and 6 months after anthracycline treatment. Patients at high risk of cardiotoxicity (cardiac troponin I concentrations in the upper tertile during chemotherapy) were randomized to standard care plus cardioprotection (combination carvedilol and candesartan therapy) or standard care alone. The primary outcome was adjusted change in left ventricular ejection fraction at 6 months. In low-risk non-randomized patients with cardiac troponin I concentrations in the lower two tertiles, we hypothesised the absence of a 6-month change in left ventricular ejection fraction (±2%).

**Results:** Between October 2017 and June 2021, 175 patients (mean age 53 years; 87% female; 71% breast cancer) were recruited. Patients randomized to cardioprotection (n=29) or standard care (n=28) had left ventricular ejection fractions of 69.4±7.4% and 69.1±6.1% at baseline and 65.7±6.6% and 64.9±5.9% 6 months after completion of chemotherapy respectively. After adjusting for age, pre-treatment left ventricular ejection fraction and planned anthracycline dose, the estimated mean difference in 6-month left ventricular ejection fraction between cardioprotection and standard care groups was -0.37% (95% confidence interval, -3.59 to 2.85%; P=0.82). In low-risk non-randomized patients, baseline and 6-month left ventricular ejection fractions were 69.3±5.7% and 66.4±6.3% respectively: estimated mean difference, 2.87% (95% confidence interval, 1.63 to 4.10%; P=0.92, not equivalent)

**Conclusions:** Combination candesartan and carvedilol therapy had no demonstrable cardioprotective effect in patients receiving anthracycline-based chemotherapy with high-risk on-treatment cardiac troponin I concentrations. Low-risk non-randomized patients had similar declines in left ventricular ejection fraction questioning the utility of routine cardiac troponin monitoring. Furthermore, the modest declines in left ventricular ejection fraction suggest that the value and clinical impact of early cardioprotection therapy needs to be better defined in patients receiving high-dose anthracycline regimes.

**REGISTRATION:** EudraCT 2017-000896-99, ISRCTN24439460

**Clinical Perspective:** *What is new?:*

- In this randomized controlled trial of patients at high risk of anthracycline cardiotoxicity, combined candesartan and carvedilol therapy did not protect against decline in 6-month left ventricular ejection fraction after completion of chemotherapy.
- Overall decline in 6-month left ventricular ejection fraction occurred irrespective of changes in cardiac troponin concentration during chemotherapy.

*What are the clinical implications?:*

- The Cardiac CARE trial findings do not support recent guideline recommendations advocating the use of cardiac troponin monitoring and early preventive neurohormonal blockade in patients at risk of anthracycline cardiotoxicity.
- Future studies should focus on factors determining transition to subsequent development of heart failure from initial mild and asymptomatic changes in cardiac function following anthracycline chemotherapy.

## INTRODUCTION

Anthracyclines are effective cytotoxic drugs that contribute to improved survival in a wide range of cancers including breast cancer and lymphoma. Anthracyclines cause dose-related cardiomyocyte injury leading to left ventricular dysfunction and heart failure.(1) Improved cancer-free survival has led to growing concern regarding the impact of cancer therapy related cardiac dysfunction (CTRCD).(2, 3) However, progression from heart muscle injury at the time of chemotherapy to the development of clinical heart failure is poorly understood and the overall utility of potentially cardioprotective treatments has not been established.(4) While extremes of age, pre-existing cardiovascular disease and cumulative anthracycline dose are risk markers for CTRCD, most anthracycline-treated patients do not develop clinically important cardiotoxicity. The degree of CTRCD observed in recent cardioprotection trials has been lower than historical studies and this may reflect a trend to the use of lower dose anthracycline regimes in high-risk patients.

International guidelines recommend cardiac troponin monitoring and cardiac imaging during and after anthracycline treatment in patients at risk of CTRCD.(5, 6) The use of potentially cardioprotective neurohormonal antagonists has been advocated for patients who are at increased risk of CTRCD because of clinical factors such as exposure to high cumulative anthracycline doses as well as patients who develop biomarker or cardiac imaging evidence of CTRCD. Cardiac troponin has been used to detect early anthracycline-induced cardiomyocyte toxicity.(7, 8) Our group and others have shown that plasma high-sensitivity cardiac troponin concentrations below the 99^th^ centile upper reference limit provide prognostic information and identify individuals with and without cardiac symptoms who are at heightened risk of cardiac events and mortality.(9)(10) Furthermore, early changes below the 99^th^ centile upper reference limit during anthracycline treatment identify those who go on to develop myocardial injury soon after chemotherapy.(11)

Set against the background of apparently declining event rates and a minority of patients developing CTRCD, previous clinical trials have adopted an unselected approach to randomization. By including patients at low risk of CTRCD, the potential to demonstrate substantial cardioprotective effects of neurohormonal blockade has been challenging.(12, 13) Additionally, most trials have used treatments that either block the renin-angiotensin system (angiotensin-converting enzyme inhibitor or angiotensin receptor antagonist) or the sympathetic nervous system (β-adrenoreceptor antagonist) but not the combination which has the most robust evidence base for improving function and survival in patients with left ventricular systolic dysfunction. The Prevention of Cardiac Dysfunction During Adjuvant Breast Cancer Therapy (PRADA) trial investigated whether anthracycline treated patients with breast cancer were protected from CTRCD with candesartan or metoprolol.(12) The overall decline in left ventricular ejection fraction (LVEF) on cardiac magnetic resonance was only 2.6 percentage points in the placebo group. Candesartan but not metoprolol had an early protective effect on LVEF decline that was not maintained on extended follow-up.(14) Consequently, it has been suggested that future trials examining cardioprotective therapy should preferentially be conducted in patients at high risk of CTRCD.(15)

The objectives of the Cardiac CARE trial were to determine whether cardiac troponin monitoring identifies patients at risk of left ventricular systolic dysfunction during anthracycline chemotherapy, and whether cardiac troponin-guided treatment with candesartan and carvedilol prevents the development of left ventricular systolic dysfunction. These objectives have immediate relevance to clinical practice and current guideline recommendations through testing a simple monitoring and threshold-guided intervention pathway that can readily be delivered within cancer treatment centers.

## METHODS

### Study design and participants

This was a multicenter prospective randomized open label blinded endpoint trial nested within an observational cohort study. Study methods and design have been described previously.(16) Eligible patients were women and men aged over 18 years, with LVEF ≥50% on baseline cardiac magnetic resonance imaging, without serious comorbidity who were scheduled for anthracycline-containing therapy for breast cancer or non-Hodgkin lymphoma. Key exclusion criteria were patients with human epidermal growth factor receptor 2 (HER2) positive breast cancer who would be scheduled for anti-HER2 therapy, lower dose anthracycline regimes (cumulative epirubicin equivalent dose <300 mg/m^2^), ongoing treatment with angiotensin receptor blockers, angiotensin-converting enzyme inhibitors or beta-blockers, hypotension or hypertension, or previous anthracycline chemotherapy (Table 1). All trial participants provided written informed consent before study procedures were conducted. The study was conducted in accordance with the Declaration of Helsinki and approved by the South-East Scotland Research Ethics Committee (17/ES/0071).

**Table 1.** Baseline characteristics of the three study groups

| Characteristic | Non-randomized<br>n = 118 | Cardioprotection<br>n = 29 | Standard care<br>n = 28 |
| --- | --- | --- | --- |
| Age (year) | 52±11 | 54±14 | 54±13 |
| Female sex | 90.7 (107) | 79.3 (23) | 78.6 (22) |
| Height (cm) | 165±8 | 167±8 | 167±8 |
| Weight (kg) | 77±17 | 71±15 | 83±17 |
| Cancer type |  |  |  |
| Breast | 78.8 (93) | 58.6 (17) | 53.6 (15) |
| Non-Hodgkin lymphoma | 21.2 (25) | 41.4 (12) | 46.4 (13) |
| Risk markers for cardiovascular disease |  |  |  |
| Smoking habit |  |  |  |
| Current smoker | 10.2 (12) | 6.9 ( 2) | 17.9 (5) |
| Ex-smoker < 1 year | 7.6 ( 9) | 6.9 ( 2) | 0 |
| Ex-smoker > 1 year | 24.6 (29) | 17.2 (5) | 17.9 (5) |
| Never smoker | 57.6 (68) | 69 (20) | 64.3 (18) |
| Diabetes mellitus |  |  |  |
| Insulin-dependent | 2.5 (3) | 0 | 0 |
| Tablet controlled | 0.8 (1) | 0 | 0 |
| Diet controlled | 0 | 0 | 0 |
| Hypertension | 8.5 (10) | 6.9 (2) | 14.3 (4) |
| Coronary disease | 2.5 (3) | 0 | 7.1(2) |
| Kidney disease | 4.2 (5) | 0 | 7.1(2) |
| Total concomitant cardiovascular medication prescriptions § | (9) | (2) | (6) |
| Cancer therapy |  |  |  |
| Cumulative anthracycline dose (mg/m<sup>2</sup>) | 424<br>[300 to 480] | 469<br>[300 to 600] | 479<br>[330 to 600] |
| 3 cycles | 29.7 (35) | 44.8 (13) | 25 (7) |
| 4 cycles | 40.7 (48) | 6.9 (2) | 25 (7) |
| 6 cycles | 29.7 (35) | 48.3 (14) | 50 (14) |
| Radiotherapy received | 71.2 (79) | 57.1 (16) | 53.6 (15) |
| Radiation target location |  |  |  |
| Left breast | 45.6 (36) | 43.8 (7) | 33.3 (5) |
| Right breast | 44.3 (35) | 37.5 (6) | 53.3 (8) |
| Both breasts | 3.8 (3) | 0 | 0 |
| Outside chest/mediastinum | 6.3 (5) | 18.8 (3) | 13.3 (2) |

§ Antihypertensive and angina medications including calcium channel blockers, thiazide and loop diuretics, nitrates and nicorandil
Mean ± standard deviation, % (n), median [interquartile range].

### Study procedures

#### Cardiac magnetic resonance

Cardiac magnetic resonance scans were conducted using steady state free-precession breath-hold cines on 1.5 and 3 Tesla magnetic resonance imaging scanners and were performed at baseline prior to commencing chemotherapy and 6 months after the final anthracycline dose. Cardiac magnetic resonance scan results were made immediately available at each site to inform ongoing clinical care. Cardiac magnetic resonance measurements for the primary and secondary outcomes including LVEF, global longitudinal and circumferential strain with cardiac magnetic resonance feature tracking, left ventricular volume and mass, and left atrial area were performed by 2 analysts in the Core Image Analysis laboratory (Edinburgh Imaging, University of Edinburgh) according to the Society for Cardiovascular Magnetic Resonance guidelines on dedicated software (CVI42 version 5.14, Circle Cardiovascular Imaging). Both analysts were independent to the research teams and blinded to scan sequence (pre or post chemotherapy) and treatment allocation.

#### Cardiac troponin monitoring

Cardiac troponin I concentrations were quantified using the ARCHITECT_*STAT*_ or ALINITY cardiac troponin assay (Abbott Laboratories, Chicago, IL) during each 3-week chemotherapy cycle. This assay has an inter-assay coefficient of variation of less than 10% at 4.7 ng/L, a limit of detection of 1.9 ng/L and a 99^th^ centile upper reference limit of 34 ng/L in men and 16 ng/L in women.(17) Patients had cardiac troponin concentrations measured prior to randomisation, before each cycle of anthracycline chemotherapy, and 2, 4 and 6 months following completion of chemotherapy.

### Study randomization and intervention

After enrolment and a baseline cardiac magnetic resonance scan, cardiac troponin concentrations were measured before each cycle of anthracycline. Patients could be randomized from cycle 2 to cycle 6. Treatment allocation was by dynamic 1:1 randomization with minimization for prognostic factors: (i) age ≥65 or <65 years, (ii) baseline LVEF ≥60% or <60%, and (iii) planned cumulative epirubicin equivalent dose 300 or >300 mg/m^2^. Cardiac troponin concentration thresholds that triggered randomization were ≥5 ng/L for cycle 2 and ≥23 ng/L for cycles 3 to 6. These thresholds were defined in a previous study as identifying patients most likely to develop myocardial injury on completion of the course of chemotherapy.(11) Non-randomized participants were treated with standard care.

The trial intervention consisted of candesartan started at 8 mg daily and increased to 16 mg and 32 mg daily, and carvedilol started at 6.25 mg twice daily, and increased to 12.5 mg and 25 mg twice daily. Combination candesartan and carvedilol were dispensed within 14 days of randomization and continued until completion or withdrawal from the study. Dose restrictions and modifications were performed according to blood pressure, heart rate and renal function.

### Outcomes

The primary efficacy outcome was adjusted change in LVEF from baseline to 6 months after the final anthracycline dose determined by cardiac magnetic resonance imaging and compared between the randomized groups. The trial also aimed to determine whether early changes in cardiac troponin concentration on anthracycline treatment identified patients at low and high risk of cardiotoxicity. To evaluate the effectiveness of cardiac troponin monitoring, the key secondary efficacy outcome was absence of change in LVEF in the low-risk non-randomized group with equivalence limits of ±2%. (18)

Other secondary efficacy outcomes included change in in global longitudinal and circumferential strain, left ventricular mass and volume, and left atria area determined by cardiac magnetic resonance imaging, and change in cardiac troponin concentration. We have previously outlined the reporting plan for clinical outcomes including cardiovascular death, new onset heart failure, additional cardiac magnetic resonance and cardiac troponin definitions of asymptomatic CTRCD, heart rate, blood pressure and safety outcomes.(15)

The analysis plan included additional exploratory comparisons across cardiotoxicity measures between low-risk non-randomized and high-risk randomized participants. These were performed to compare the magnitude of plasma cardiac troponin concentration increase in randomized and non-randomized participants, and to determine how closely the trial protocol identified a population at risk of cardiotoxicity.

The following investigational medicinal product safety outcomes were compared between treatment groups; hypotension (systolic blood pressure < 90 mmHg), bradycardia (heart rate < 50 bpm), hyperkalaemia (≥ 5.0 mmol/L), worsening renal function, acute kidney injury, fatigue and new atrial fibrillation.

### Statistical analysis

Descriptive data are presented as number (percent), mean ± standard deviation or median [interquartile range].

We planned to randomize at least 33% of participants using the previously defined cardiac troponin thresholds.(11,15) We assumed that this threshold would select all participants developing clinically important reductions in LVEF. Assuming a standard deviation of 5% in LVEF,(4) we required 23 participants per group to detect a 5% change in LVEF at 90% power and two-sided p<0.05. This effect size was used in the PRADA Study.(19) Allowing for 1 in 6 participants with missing data, the total randomized trial sample size was 56. Assuming a third of participants would be randomized into the trial, the total observational study cohort size was at least 168 participants. The primary analysis was adjusted for the binary fixed effects: age at consent ≥65 or <65 years; baseline LVEF ≥60% or <60% and planned cumulative epirubicin equivalent dose =300 mg/m^2^ or >300 mg/m^2^. This was an intention-to-treat analysis, and treatment effect was expressed by a point mean difference estimate and its 95% confidence interval (CI). This approach was also used for the efficacy of candesartan and carvedilol treatment on secondary outcomes from cardiac magnetic resonance and cardiac troponin concentrations at 2 months. We endeavoured to keep missing values to a minimum, and the primary analysis was a complete case analysis. If there were sufficient missing data to cause concern, multiple imputation was to be used as a sensitivity analysis, but the primary outcome did not have sufficient missing data to cause concern (<10%), and multiple imputation was not necessary.

To assess the effectiveness of cardiac troponin monitoring, we measured change in LVEF in the low-risk non-randomized group defining an absence of change in LVEF as being within ±2% of the baseline measurement. At 90% power and two-sided p<0.05 and assuming a similar standard deviation of 5%, paired cardiac magnetic resonance imaging scans would be required in 68 non-randomized participants.

Other secondary efficacy and safety outcomes were analysed as appropriate with linear regression for continuous outcomes, logistic regression for binary outcomes, and Cox proportional hazards for survival analysis, adjusted as per the primary analysis. A full Statistical Analysis Plan was finalized before database lock.

## RESULTS

### Study population

Between 4^th^ October 2017 and 30 ^th^ June 2021, 424 patients were approached across 7 centers in the United Kingdom (Figure 1). Of these, 191 (45.0%) patients consented to participation and 16 patients were excluded. From 175 patients included in the observational cohort, 57 (32.6%) were randomized into the trial: 50 (87.7%) were randomized at anthracycline treatment cycle 2, 4 (7.0%) at cycle 5 and 3 (5.3%) at cycle 6.

**Figure 1.**
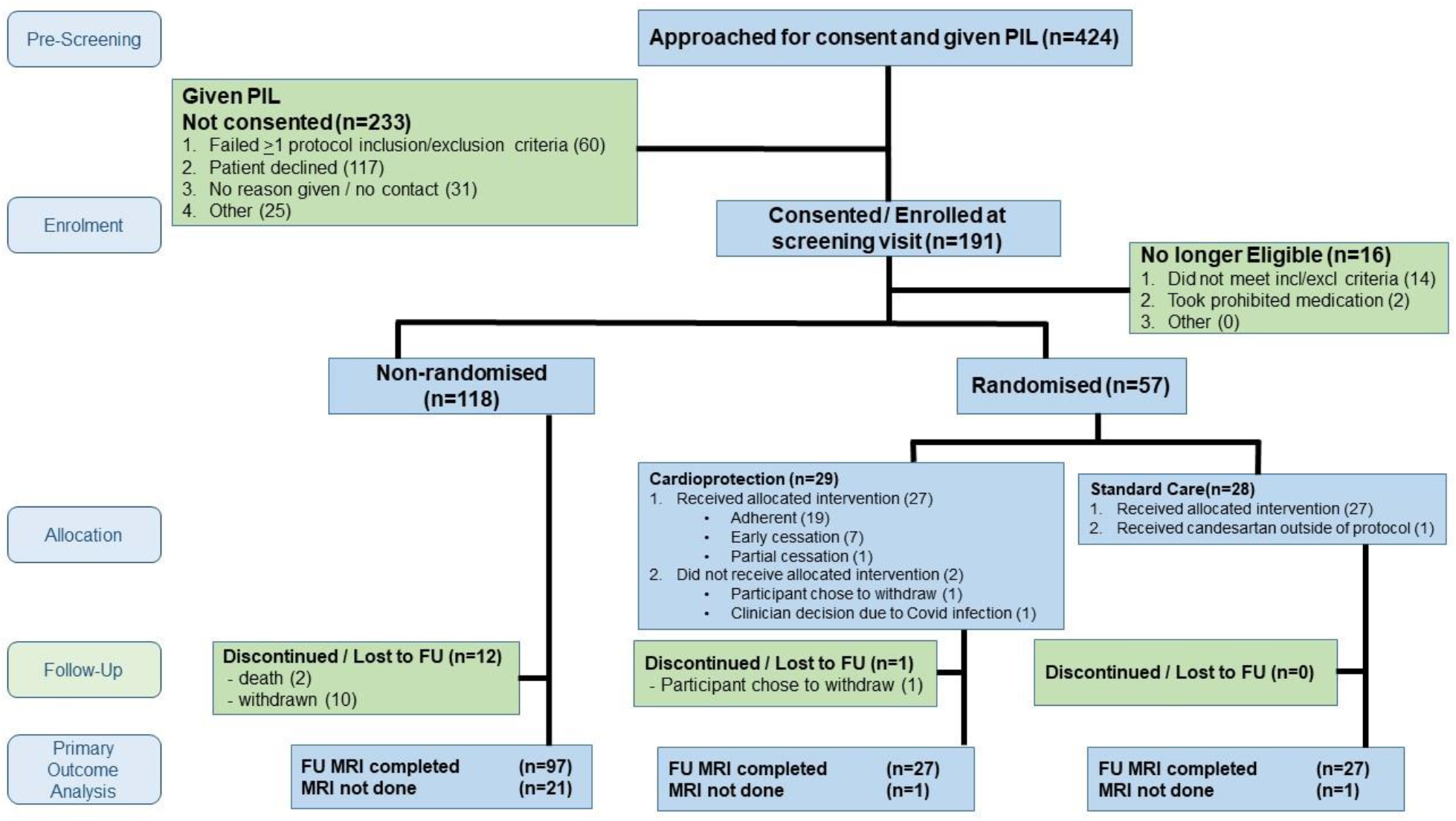
Cardiac CARE Consolidated Standards of Reporting Trials Diagram. Cardiac CARE screening and randomization. The number of patients at the screening, enrolment, randomization, 6 month follow up and primary outcome analysis are shown. Withdrawals and adherence are indicated. FU; follow up, MRI; magnetic resonance imaging; IMP investigational medicinal product; PIL patient information leaflet

The study population was predominantly composed of women with breast cancer (Table 1). Patients with non-Hodgkin lymphoma were proportionately more likely to be randomized than patients with breast cancer. Cardiovascular risk factors and concomitant cardiovascular medications were uncommon across all groups. Mean anthracycline dose was higher in the randomized groups. Radiotherapy was more commonly prescribed in the non-randomized group (71.2%) compared to the cardioprotection (57.1%) and standard care groups (53.6%).

### Compliance with treatment allocation

Twenty patients (68.9%) were adherent to cardioprotection treatment at 6 months although 1 stopped candesartan within 2 months and continued with carvedilol alone. Two patients (6.9%) randomized to cardioprotection did not receive any medication owing to intercurrent illness and COVID-19 infection. A further 7 (24.1%) participants stopped both cardioprotection drugs within 2 months owing to symptoms of light headedness and dizziness possibly related to low blood pressure. Blood pressure and heart rate were lower in the cardioprotection treatment group at 6 months (online table 2). On post-hoc analysis, there was a greater reduction in heart rate at 6 months in the cardio-protection group: estimated mean difference –11 bpm (95% CI, -18 to -4 bpm; P = 0.003). Blood pressure appeared to be lower in the cardioprotection group but changes in systolic (−7 mmHg (95% CI, -17 to +2.0 mmHg; P = 0.12) and diastolic (−6 mmHg (95% CI, -12 to +0.2 mmHg; P= 0.06) pressures were not statistically significant.

**Table 2.** Participant hemodynamic measures at baseline, 2, 4 and 6 months following completion of chemotherapy.

|  | **Baseline** |  | **2 months** |  | **4 months** |  | **6 months** |  |
| --- | --- | --- | --- | --- | --- | --- | --- | --- |
| Parameter | Cardio-protection | Standard care | Cardio-protection | Standard care | Cardio-protection | Standard care | Cardio-protection | Standard care |
| HR bpm | 77 ± 12 | 82 ± 13 | 80 ± 12 | 84 ± 14 | 72 ± 11 | 79 ± 10 | 74 ± 9 | 85 ± 13 |
| SBP mmHg | 131 ± 17 | 132 ± 18 | 120 ± 22 | 132 ± 17 | 121 ± 14 | 131 ± 18 | 119 ± 17 | 128 ± 15 |
| DBP mmHg | 80 ± 12 | 80 ± 11 | 68 ± 11 | 81 ± 9 | 75 ± 9 | 80 ± 9 | 73 ± 11 | 79 ± 9 |

Mean ± standard deviation
Where: HR, heart rate; SBP, systolic blood pressure; DBP, diastolic blood pressure

### Primary and key secondary outcomes

Patients randomized to cardioprotection or standard care had LVEFs of 69.4±7.4% and 69.1±6.1% at baseline and 65.7±6.6% and 64.9±5.9%, 6 months after completion of chemotherapy respectively (online Table 3). After adjusting for age, pre-treatment LVEF and planned anthracycline dose, there was no change in the estimated mean difference in 6-month LVEF between cardioprotection and standard care groups (−0.37 percentage points, 95% CI, -3.59 to 2.85; P=0.82, Central Figure 2A).

**Table 3.**
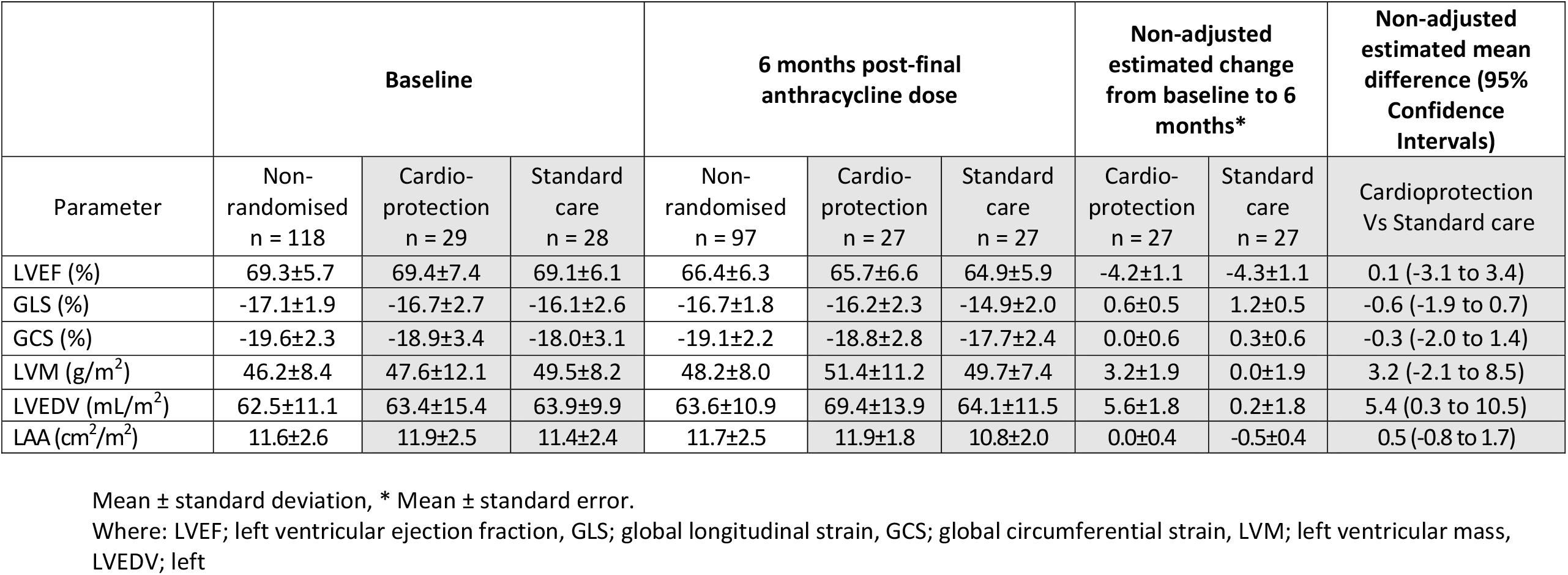
Participant cardiac MRI measures at baseline and 6 months post anthracycline. Non-adjusted estimated mean difference between randomised groups at 6 months.

|  | Baseline |  |  | 6 months post-final anthracycline dose |  |  | Non-adjusted estimated change from baseline to 6 months* |  | Non-adjusted estimated mean difference (95% Confidence Intervals) |
| --- | --- | --- | --- | --- | --- | --- | --- | --- | --- |
| Parameter | Non-randomised<br>n = 118 | Cardio-protection<br>n = 29 | Standard care<br>n = 28 | Non-randomised<br>n = 97 | Cardio-protection<br>n = 27 | Standard care<br>n = 27 | Cardio-protection<br>n = 27 | Standard care<br>n = 27 | Cardioprotection Vs Standard care |
| LVEF (%) | 69.3±5.7 | 69.4±7.4 | 69.1±6.1 | 66.4±6.3 | 65.7±6.6 | 64.9±5.9 | -4.2±1.1 | -4.3±1.1 | 0.1 (-3.1 to 3.4) |
| GLS (%) | -17.1±1.9 | -16.7±2.7 | -16.1±2.6 | -16.7±1.8 | -16.2±2.3 | -14.9±2.0 | 0.6±0.5 | 1.2±0.5 | -0.6 (-1.9 to 0.7) |
| GCS (%) | -19.6±2.3 | -18.9±3.4 | -18.0±3.1 | -19.1±2.2 | -18.8±2.8 | -17.7±2.4 | 0.0±0.6 | 0.3±0.6 | -0.3 (-2.0 to 1.4) |
| LVM (g/m<sup>2</sup>) | 46.2±8.4 | 47.6±12.1 | 49.5±8.2 | 48.2±8.0 | 51.4±11.2 | 49.7±7.4 | 3.2±1.9 | 0.0±1.9 | 3.2 (-2.1 to 8.5) |
| LVEDV (mL/m<sup>2</sup>) | 62.5±11.1 | 63.4±15.4 | 63.9±9.9 | 63.6±10.9 | 69.4±13.9 | 64.1±11.5 | 5.6±1.8 | 0.2±1.8 | 5.4 (0.3 to 10.5) |
| LAA (cm<sup>2</sup>/m<sup>2</sup>) | 11.6±2.6 | 11.9±2.5 | 11.4±2.4 | 11.7±2.5 | 11.9±1.8 | 10.8±2.0 | 0.0±0.4 | -0.5±0.4 | 0.5 (-0.8 to 1.7) |

Mean ± standard deviation, \* Mean ± standard error.
Where: LVEF; left ventricular ejection fraction, GLS; global longitudinal strain, GCS; global circumferential strain, LVM; left ventricular mass, LVEDV; left

**Figure 2A.**
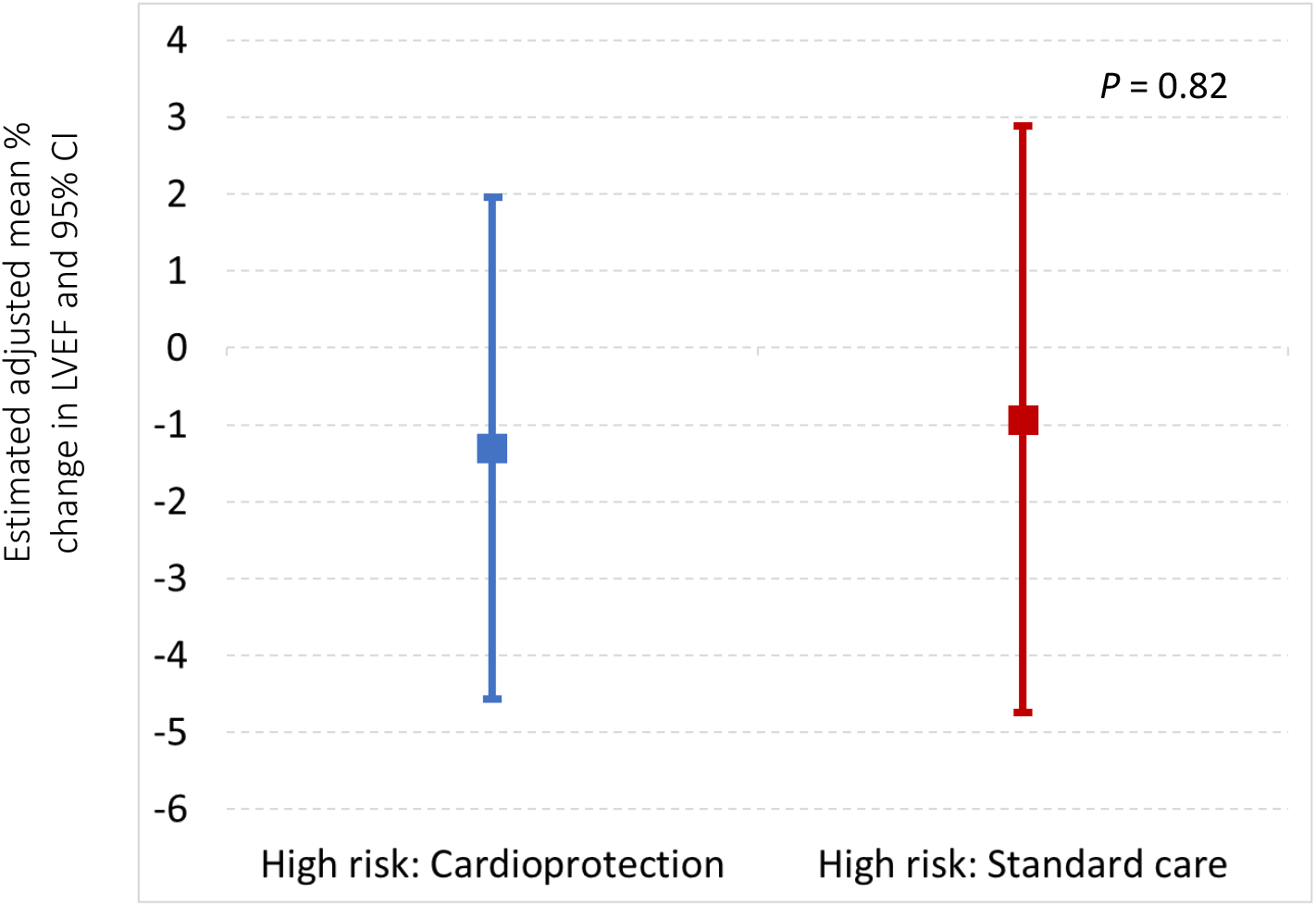
Central Illustration. Primary outcome: Change in left ventricular ejection fraction (LVEF) adjusted for prognostic factors^1^ LVEF; left ventricular ejection fraction, CI confidence intervals 1. Age ≥ 65 or < 65 years, LVEF at baseline ≥ 60% or < 60%, cumulative epirubicin dose = 300 mg/m^2^ or > 300 mg/m^2^

**Figure 2B.**
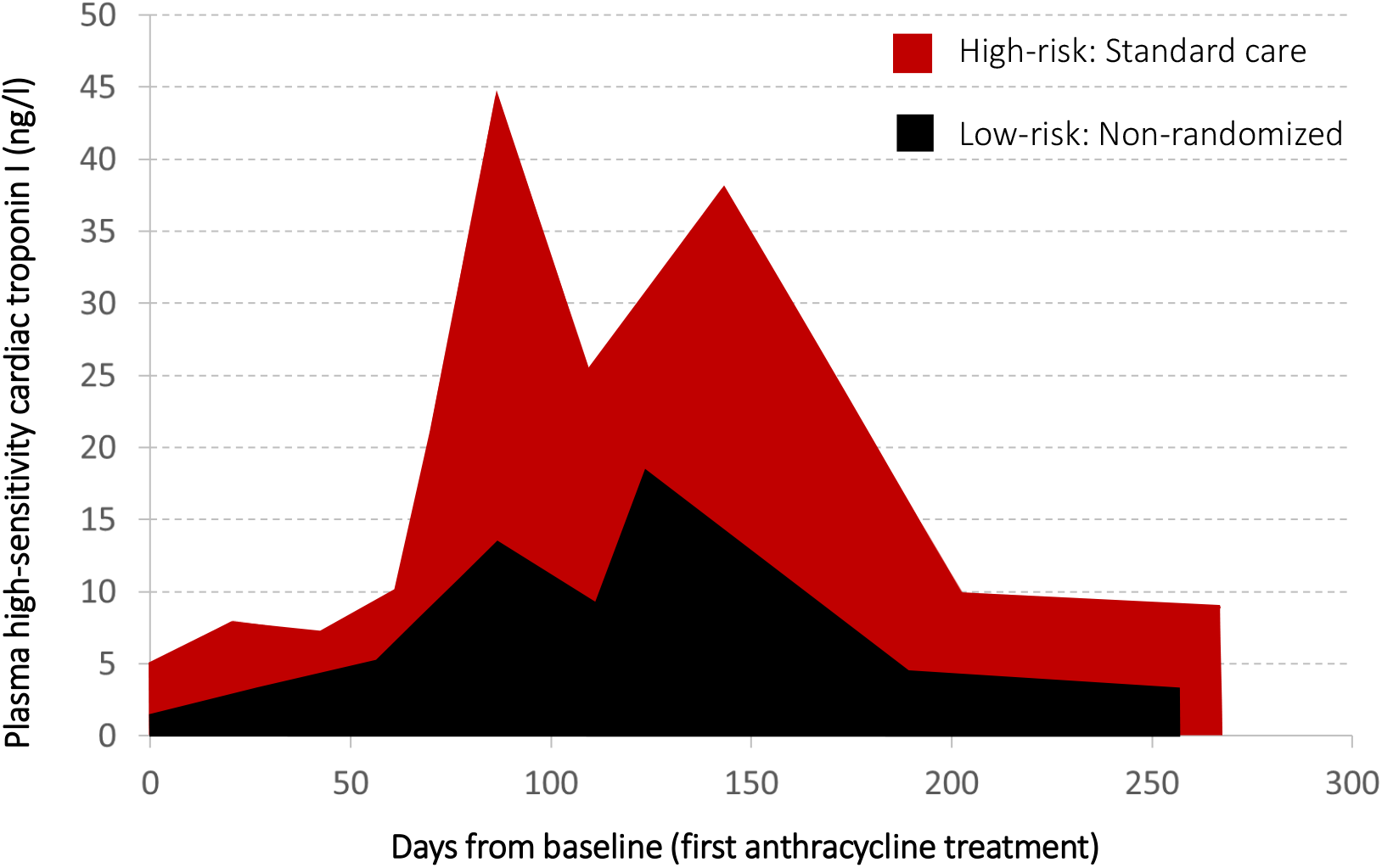
Central Illustration. Average high sensitivity cardiac troponin I (hs-cTnI) concentrations^1^ in high risk (randomized-to-standard-care) and low risk non-randomized groups. 1. Abbot ARCHITECHT hs-cTnI assay. Sex specific 99^th^ centile URL; <16 ng/L (female), <34 ng/L (male)

In non-randomized patients, the baseline and 6-month LVEFs were 69.3±5.7% and 66.4±6.3%, respectively. The estimated non-adjusted mean difference was 2.87% and its 95% CI was 1.63% to 4.10%. Hence the main secondary objective of demonstrating no change in LVEF with equivalence of ±2% was not met (P=0.92).

In post-hoc sensitivity analyses, we assessed the per protocol primary efficacy outcome between randomized groups. There was no difference in outcome when only the 19 cardioprotection patients who were fully adherent with treatment allocation were included. The estimated mean difference in the change in 6-month LVEF between cardioprotection and standard care groups was -0.7 percentage points (95% CI, -4.3 to 2.9; P=0.70).

### Secondary outcomes

Cardiac troponin I concentrations progressively increased in all groups during chemotherapy (online Table 4). By design, cardiac troponin concentrations were higher in the randomized groups (Central Figure 2B and online Table 5). Adjusted estimated mean change (± standard error) in cardiac troponin concentration from baseline to 2 months post chemotherapy in the cardioprotection and standard care groups was 27.3±7.4 and 28.8±8.8 ng/L respectively. The adjusted estimated mean difference was -1.55 ng/L (95% CI -17.56 to 14.45 ng/L; P=0.85). A difference between cardioprotection and standard care groups was observed for the secondary outcome of adjusted left ventricular end diastolic volume indexed for body surface area (Table 6). However, there were no differences for global longitudinal and circumferential strain, left ventricular mass and left atrial area.

**Table 4.** Adjusted change in cardiac magnetic resonance measures from baseline to 6 months after final anthracycline dose.

|  | **Adjusted † estimated change from baseline to 6 months** |  |  |  |
| --- | --- | --- | --- | --- |
| Parameters | Cardioprotection<br>n = 27 | Standard care<br>n = 27 | Estimated mean difference (CI) | *p* - value |
| LVEF(%)§ | -1.3 (1.6) | -0.9 (1.9) | -0.4 (-3.6 to 2.8) | 0.82 |
| GLS (%) | 0.1 (0.7) | 0.5 (0.8) | -0.4 (-1.8 to 1.0) | 0.59 |
| GCS (%) | -2.0 (0.8) | -2.2 (1.0) | 0.2 (-1.5 to 1.8) | 0.84 |
| LVM g/m<sup>2</sup> | 1.9 (2.8) | -1.9 (3.3) | 3.8 (-1.9 to 9.4) | 0.18 |
| LVEDV ml/m<sup>2</sup> | 3.4 (2.7) | -2.6 (3.2) | 6.0 (0.6 to 11.4) | 0.03 |
| LAA cm<sup>2</sup>/m<sup>2</sup> | 0.4 (0.7) | 0.1 (0.8) | 0.3 (-1.0 to 1.6) | 0.65 |

† Outcome adjusted for age at consent ≥ 65 or < 65 years, LVEF at baseline ≥ 60% or < 60% and planned cumulative epirubicin equivalent dose =300 mg/m<sup>2</sup> or >300 mg/m<sup>2</sup>
§ Primary Outcome
Where: LVEF; left ventricular ejection fraction, GLS; global longitudinal strain, GCS; global circumferential strain, LVM; left ventricular mass, LVEDV; left ventricular end diastolic volume, LAA; left atrial area.

**Table 5.** Participant clinical outcomes and measures of chemotherapy-related cardiac dysfunction.

| Outcome<br>% (no.) | Non-randomised<br>n = 118 | Cardioprotection<br>n = 28 | Standard care<br>n = 29 |
| --- | --- | --- | --- |
| Cardiovascular death | 0 | 0 | 0 |
| Any new heart failure | 0 | 0 | 3.6 (1) |
| Any new atrial fibrillation | 0 | 0 | 0 |
| Any $\geq 10$ % point fall AND<br>absolute LVEF fall below 50% | 0 | 0 | 0 |
| Any fall in LVEF below 50% | 0 | 0 | 0 |
| GLS fall > 15% | 6.5 (6) | 14.8 (4) | 3.7 (1) |
| Chronic myocardial injury | 32.1 (34) | 35.7 (10) | 60 (15) |
| Any hs-cTnI concentration > 80<br>ng/L | 0 | 10.3 (3) | 17.9 (5) |

Where: LVEF; left ventricular ejection fraction, GLS; global longitudinal strain, hs-cTnI; high sensitivity cardiac troponin I

**Table 6.** Patient adverse event reporting. Number or percentage (number)

| Parameter | Non-randomised<br>n = 118 | Cardioprotection<br>n = 28 | Standard care<br>n = 29 |
| --- | --- | --- | --- |
| Number of adverse events | 18 | 32 | 3 |
| Patients with any adverse event | 12.7 (15) | 71.4 (20) | 10.3 (3) |
| Serious adverse events | 61.1 (11/18) | 37.5 (12/32) | 66.7 (2/3) |
| Adverse event possibly related to the investigational medicinal product | 5.6 (1/18) | 62.5 (20/32) | 0 |

Exploratory comparisons were conducted between the high-risk standard care group and low-risk non-randomized group. The non-adjusted mean change in LVEF in the low-risk non-randomized participants was -2.9±6.1% as compared to -4.33±4.4% in the high-risk standard care group and they were similar (mean difference -1.46%, 95% CI -3.96 to 1.03, P=0.25).

The adjusted estimated mean difference in LVEF decline between these two groups was similar. Adjusted estimated mean difference in all cardiac magnetic resonance measures of cardiotoxicity between the high-risk standard care group and low-risk non-randomized groups were similar (online Table 7).

### Clinical outcomes

There were no cardiovascular deaths or incident episodes of atrial fibrillation recorded during the trial (Table 8). One patient randomized to the standard care group developed heart failure and received treatment for this including candesartan. This patient’s LVEF recovered on the 6-month cardiac magnetic resonance scan. No patients met the CTRCD criterion of a 10-percentage-point LVEF fall or a fall to an absolute LVEF below 50%.

Similarly, the CTRCD criterion of greater than 15% fall in global longitudinal strain was uncommon across groups. Chronic myocardial injury defined as an elevated cardiac troponin concentration about the 99^th^ centile upper reference limit 2 months after completion of chemotherapy was common and similar in non-randomized (32.1%) and cardio-protection (35.7%) groups. The proportion with chronic myocardial injury was higher (60%) in the standard care treatment group. Recordings of high (> 80 ng/L) cardiac troponin concentrations were confined to randomized groups.

### Safety outcomes

Adverse events were more commonly reported in the cardioprotection group with 71.4% of patients having at least one adverse event comparted to 10.3% standard care patients and 12.7% non-randomized patients (Table 9). During the trial there were 25 reportable serious adverse events with 12 in the cardioprotection group compared to 2 in the standard care group. Most adverse events in the cardioprotection group were possibly related to the investigational medicinal products with dizziness and syncope listed in 17 out 20 possibly related adverse events and hypotension, palpitation and venous thrombo-embolism listed for the remaining adverse events with possible causal link with the investigational medicinal product. By contrast to adverse event reporting, there was no signal of harm related to investigational medicinal product prescription from safety reporting. There were no patients with protocol defined hypotension or bradycardia at study visits following completion of chemotherapy. Hyperkalaemia at any point after randomisation occurred in 10.3 % of non-randomized patients and was more common in the cardioprotection (20.7%) group and standard care (17.9%) groups. Worsening renal function at any point beyond baseline occurred in 2.7%, 6.9% and 7.1% of non-randomized, cardioprotection and standard care groups, respectively. Fatigue was reported by 12.1%, 3.4% and 25.0% of non-randomized, cardio-protection and standard care groups, respectively.

## DISCUSSION

We found no strong evidence that early cardioprotection therapy with combined candesartan and carvedilol therapy prevented 6-month decline in LVEF in patients with breast cancer or non-Hodgkin lymphoma. This was despite enrichment by only randomizing patients with high cardiac troponin concentrations and clear evidence of pharmacological effect with changes in heart rate and left ventricular end diastolic volume in the cardioprotection group. Moreover, LVEF decline was similar in low-risk non-randomized and high-risk randomized patient groups despite substantial differences in cardiac troponin concentrations during anthracycline treatment. Overall, our findings question the benefit of guidelines that advocate on treatment cardiac troponin monitoring to identify patients at risk of anthracycline CTRCD and early intervention with cardioprotection therapy in patients with the highest levels of cardiac troponin.

Key strengths of this multicenter trial include cardiac magnetic resonance imaging with core laboratory quantification and use of the same high sensitivity cardiac troponin I assay across all sites to provide precise and standardised measures of myocardial injury. Recruitment and randomisation goals were exceeded and data completeness for the primary and main secondary outcomes was excellent despite the challenge of the Covid-19 pandemic that interrupted trial recruitment for 6 months.

The decline in LVEF at 6 months was smaller than previous studies using echocardiographic monitoring (20, 21) but comparable to recent multicenter studies enrolling similar patient populations using cardiac magnetic resonance.(14, 22) We observed small deteriorations in both global and longitudinal strain across all groups. There was no difference in these early markers of ventricular dysfunction between randomized groups and the cardiotoxicity threshold of > 15% relative fall in strain was uncommon at 6 months. The increase in left ventricular end-diastolic volume in the cardioprotection group was likely related to the impact of beta-blockers slowing heart rate with consequent increased filling and stroke volume.

Candesartan and carvedilol therapy did not reduce cardiac troponin concentration change from baseline to 2 months post-chemotherapy. We identified this as a time point to examine for a treatment effect when cardiac troponin concentrations might be expected to remain elevated after completion of chemotherapy, and when patients randomized to cardioprotection would have received therapy for at least 2 months. Participants with the highest concentrations of cardiac troponin (any measurement > 80 ng/L) were confined to randomized groups and chronic myocardial injury, defined as a persistent elevation in cardiac troponin above the 99^th^ centile upper reference limit, 2 months after chemotherapy was common across all 3 groups. We believe this is the first time this persistent signal of myocardial injury, present in 60% of those randomized to standard care group, has been demonstrated in a large population of patients receiving anthracycline cardiotoxicity.

Combined therapy with candesartan and carvedilol, initiated during chemotherapy, was associated with adverse effects and 31% of patients stopped or did not start cardioprotection therapy within 2 months of randomisation. Symptoms possibly related to cardioprotection medication, such as dizziness, were frequently listed in adverse event reporting as the reason for early cessation. By contrast, the rate of non-adherence was lower in the PRADA study with 7% of patients assigned to the combined metoprolol and candesartan therapy arm discontinuing medication,(12) and this may reflect use of placebo control in this study. As in PRADA, we found no evidence for a signal of excess harm related to cardioprotection therapy in the protocol specified investigational medicinal product safety outcomes. Fatigue was more common in those randomized to standard care. This result was unexpected given that fatigue is a side effect commonly attributed to beta-blockade.

### Limitations

Several patients discontinued cardio-protection medication within 2 months of randomization and this may have had some influence on treatment effect. The trial was powered to detect a 5%-point difference in LVEF between randomized groups and there was no evidence of greater or less LVEF decline with cardioprotection. However, we cannot exclude a small treatment effect although the upper boundary of the 95% confidence interval for the primary endpoint was 2.85%.

## Conclusions

Six months after completion of anthracycline chemotherapy, combined candesartan and carvedilol did not prevent a small decline in LVEF or impact on other secondary CTRCD measures. Our results are in accordance with recent trials investigating neurohormonal blockade in anthracycline treated patients.(13, 14) The trial protocol successfully identified participants who developed high on treatment cardiac troponin concentrations for randomization, but despite being considered high risk, the degree of CTRCD in the randomized group was mild and not demonstrably different from low-risk non-randomized participants. Cardioprotection therapy was poorly tolerated by some participants leading to drug-related side effects and discontinuations. The small decline in LVEF observed in Cardiac CARE will not have immediate clinical implications for individual patients. Applied across a population, this LVEF decline may confer a general increased risk of future cardiac dysfunction and heart failure. Our findings show that the benefit of targeted combined cardioprotection therapy with candesartan and carvedilol is uncertain and not well tolerated. Future research should be directed at understanding factors determining evolution of late cardiac dysfunction in this patient population with more prolonged monitoring, long-term imaging and clinical follow up.

## Data Availability

Additional data and the full study protocol are availablenfrom the corresponding author upon reasonable request.

## Disclaimer

The views expressed in this publication are those of the author and not necessarily those of the MRC, NHS, NIHR or the Department of Health.

*The EME Programme is funded by the MRC and NIHR, with contributions from the CSO in Scotland and NISCHR in Wales and the HSC R&D Division, Public Health Agency in Northern Ireland

## Acknowledgements

The authors would like to thank all the patients, site staff, the image analysis team, the trial steering committee, the data monitoring committee, and the patient advisory group. We thank Edinburgh Clinical Trials Unit staff for their involvement. The University of Edinburgh and the Lothian Health Board are co-sponsors. We thank the NIHR and British Heart Foundation for funding the trial and for their constant support and advice. We acknowledge the support of the NIHR clinical research network. MRI analysis was expertly performed at the Edinburgh Imaging Facility, University of Edinburgh. The following groups all contributed substantially to the trial.

## Trial steering committee

Prof Stephen Evans (Chairperson, Professor of Pharmacoepidemiology at London School of Hygiene and Tropical Medicine, UK), Prof Torbjørn Omland (Professor of Medicine at Akershus, Oslo, Norway), Prof Abigail Marks (patient representative), Dr Gordon Urquhart (Consultant Medical Oncologist at Aberdeen Royal Infirmary, UK), Prof Russell Petty (Chair of Medical Oncology at Ninewells Hospital and Medical School, Dundee, UK).

## Data monitoring committee

Prof Helena Earl (Chairperson prior to retirement, Consultant Medical Oncologist at Addenbrooke’s Hospital, UK), Dr Mark Francis (Chairperson, Consultant Cardiologist at NHS Fife, UK), Dr Ellen Copson (University of Southampton, UK), Ms Evie Gardner (Statistician Northern Ireland Clinical Trial Unit, UK), Dr Dominic Culligan (Consultant haematologist at Aberdeen Royal Infirmary and The University of Aberdeen, UK), MS Helen Mossop (Biostatistics Research Group within the Population Health Sciences Institute at Newcastle University, UK)

## Trial management group

the Edinburgh Clinical Trials Unit staff: Anna Foster (trial management support) Hannah Rickman (trial management support) Garry Milne (database programmer), Lynsey Milne (data management), Linda Williams (unmasked statistician).

## Edinburgh Imaging Facility

Tom MacGillivray (Senior Research Fellow), Scott Semple (Reader in Medical Physics), Annette Cooper (lead radiographer)

## Contribution statement

PAH, PH, OO, MM, SL, DEN, NLM and NL conceived the study and its design. AR had access to the data and performed the analysis. PAH, SL, AR, PH, OO, DEN, NLM and NL interpreted the data and drafted the manuscript. All authors revised the manuscript critically for important intellectual content and provided their final approval of the version to be published. All authors are accountable for the work.

